# Structural and spatial dynamics of mosquito-arbovirus associations in México (2007 to 2025): A systematic review and quantitative synthesis

**DOI:** 10.64898/2026.04.24.26351690

**Authors:** Ayokunle D. Obayomi, Jonathan P. Cisneros, Marvellous Asubiojo, Omar Domínguez-Acosta, Temidayo O. Elufisan, Ma Isabel Salazar

## Abstract

**Background:** Mosquito-borne arboviruses present persistent public health threats in México. Multiple vector species are often considered to influence the local transmission of arboviral diseases; however, the structure and spatial dynamics of mosquito-arbovirus associations are unknown.

**Methods:** We conducted a systematic review to synthesize research investigating natural arboviral infections across mosquito taxa. PRISMA-guided search was done in PubMed, Scopus, Web of Science and Google Scholar, resulting in 46 included articles from 2007 to 2025. To delineate mosquito-arbovirus associations, spatial autocorrelation, bipartite network analysis, generalized linear mixed models (GLMMs) and comparative analysis of infection across sex and life stages were used to resolve spatial dynamics, species-specific viral detection and maintenance profiles.

**Results:** Minimum Infection Rate (MIR) revealed significant positive spatial autocorrelation (Global Moran’s I = 0.139; p = 0.016), indicating structured but diffuse spatial patterns (Local Moran’s I = 0.092; p = 0.045). Sampling intensity negatively correlated with the MIR (Spearman ρ = −0.680, p < 0.001), indicating that sampling effort did not obscure the spatial structure. Elevated values were observed in a few municipalities across south-central and southeastern Mexico, where vegetation and land use interface. Network analysis (connectance = 0.40) and GLMM characterized an *Aedes*-centered network with broader taxonomic patterns driven by ZIKV; detection was significantly higher in *Culex quinquefasciatus* (OR = 1.88, p < 0.001) relative to *Aedes aegypti*. DENV detection patterns contrasted with other key viruses; no significant differences in transmission modes (χ² = 1.01, p = 0.315), suggesting a distinct maintenance profile.

**Conclusions:** This review unveils spatially diffuse and virus-specific detection patterns across heterogeneous communities in México, findings that transcend *Aedes*-centric frameworks. These resolutions provide an evidence-based baseline that encourages an integrated, community-scale approach in regional surveillance programs.

**AUTHOR SUMMARY:** Mosquito-borne arboviruses continue to threaten human and animal health across México. Previous research has investigated arboviral infections and mosquito fauna as separate entities, but not their associations. This systematic review aims to resolve the spatial structure and multi-species dynamics of mosquito–arbovirus associations. We synthesized a long-term dataset of mosquito-borne virus surveillance from academic articles. Spatial models determined if localized detections were random and/or clustered with hotspots. Network and mixed models estimated the likelihood of positive arbovirus detection in mosquito species and a comparison test quantified the influence of vertical transmission on viral maintenance. Arboviral infection rates showed a diffuse spatial pattern with elevated values across urban and interface settings. *Aedes aegypti* shaped the mosquito–arbovirus network as expected; however, *Culex quinquefasciatus* showed significantly higher odds of ZIKV detection, supporting broader surveillance. These findings clarify species-specific detection and maintenance profiles across mosquito taxa, informing improved and targeted control strategies for arboviral diseases in México.

## INTRODUCTION

Mosquito-borne arboviruses remain a public health concern as the main etiological agents of vector-borne diseases in the Americas [1]. More than 17% of infectious diseases, which cause 700,000 deaths each year, are vector-borne [2]. Mosquito-borne viruses are primarily represented by dengue virus (DENV), yellow fever virus (YFV), West Nile virus (WNV), Zika virus (ZIKV) and Chikungunya virus (CHIKV) [3]. Notably, dengue caused by DENV is the most prevalent, accounting for approximately 90 million cases and 40,000 deaths annually [2]. Arboviral transmission features host-vector-virus associations with mosquitoes supporting maintenance across different ecosystems [4, 5].

México represents quintessential biodiversity in the Americas, hosting 23 vector and 233 non-vector mosquito species with documented arbovirus circulation in different ecosystems. This biodiversity is distributed across ecological transition zones covering peri-urban, agricultural and fragmented forest landscapes [5]. Recent Pan-American alerts on yellow fever noted sylvatic expansions through forest edges and anthropogenic interfaces beyond historical hotspots [6].

As the body of entomo-virological evidence has matured, arbovirus detections have been reported from different mosquito species across diverse ecological regions [7, 8], indicating a shift towards a broader surveillance framework. Nevertheless, fragmented research with inconsistent metrics focuses on *Aedes (Ae.) aegypti* [9, 10], leaving secondary detections debated as incidental spillover [8] and limiting national-scale understanding of how mosquito–arbovirus detections are structured across taxa and landscapes.

Previous syntheses have described the mosquito fauna and its distribution to provide entomological context [5, 11], prioritized epidemiological history [1] and identified urban hotspots of key arboviruses through clinical cases [12]. However, these efforts were often restricted to taxa or regions, without integrating virus detection data across mosquito taxa, landscapes and surveillance methods.

Here, we synthesize evidence from published articles (2007-2025) investigating natural arboviral infections in mosquitoes to resolve arbovirus detection across space and mosquito taxa in México. Specifically, this study aims to characterize spatial structure, taxonomic detection patterns and the mosquito-arbovirus associations within an analytical framework. We further evaluate evidence of vertical transmission in primary vectors for key arboviruses.

## METHODS

### Electronic and manual search strategy

This systematic review followed the Preferred Reporting Items for Systematic Reviews and Meta-analyses (PRISMA) guidelines [13]. Electronic database searches in PubMed, Scopus, and Web of Science yielded 1172 articles, and manual screening in Google Scholar returned 200 additional articles; all were studies conducted in México from 1 January 2007 to 30 November 2025. The search used the following combination of relevant keywords: ((“Dengue” OR “Zika” OR “Chikungunya” OR “Yellow Fever” OR “West Nile” OR “Flavivirus” OR “Alphavirus” OR “Bunyavirus” OR “Mosquito-borne virus”) AND (México) AND (“Human” OR “Animal” OR “Mosquito”) AND (detection OR surveillance OR prevalence)). Studies including human or animal infection data were retained where mosquito-based detection was also reported, and only the entomological component was extracted for analysis. Importation of references and deduplication were done in Zotero version 6.0.36 (Fairfax, VA).

### Inclusion and exclusion criteria

The inclusion criteria were: (1) studies conducted in México, and (2) original research investigating the natural infection of mosquito taxa with vertebrate-pathogenic arboviruses. We excluded studies if they: (1) were not conducted in Mexico, (2) were not primary research with mosquito-arbovirus associations, or (3) lacked sufficient methodological detail. Titles, abstracts, and full-texts were screened independently by two authors and unresolved disagreements by a third reviewer, with detailed exclusion categories in the flow diagram (Fig. 1).

**Fig 1.**
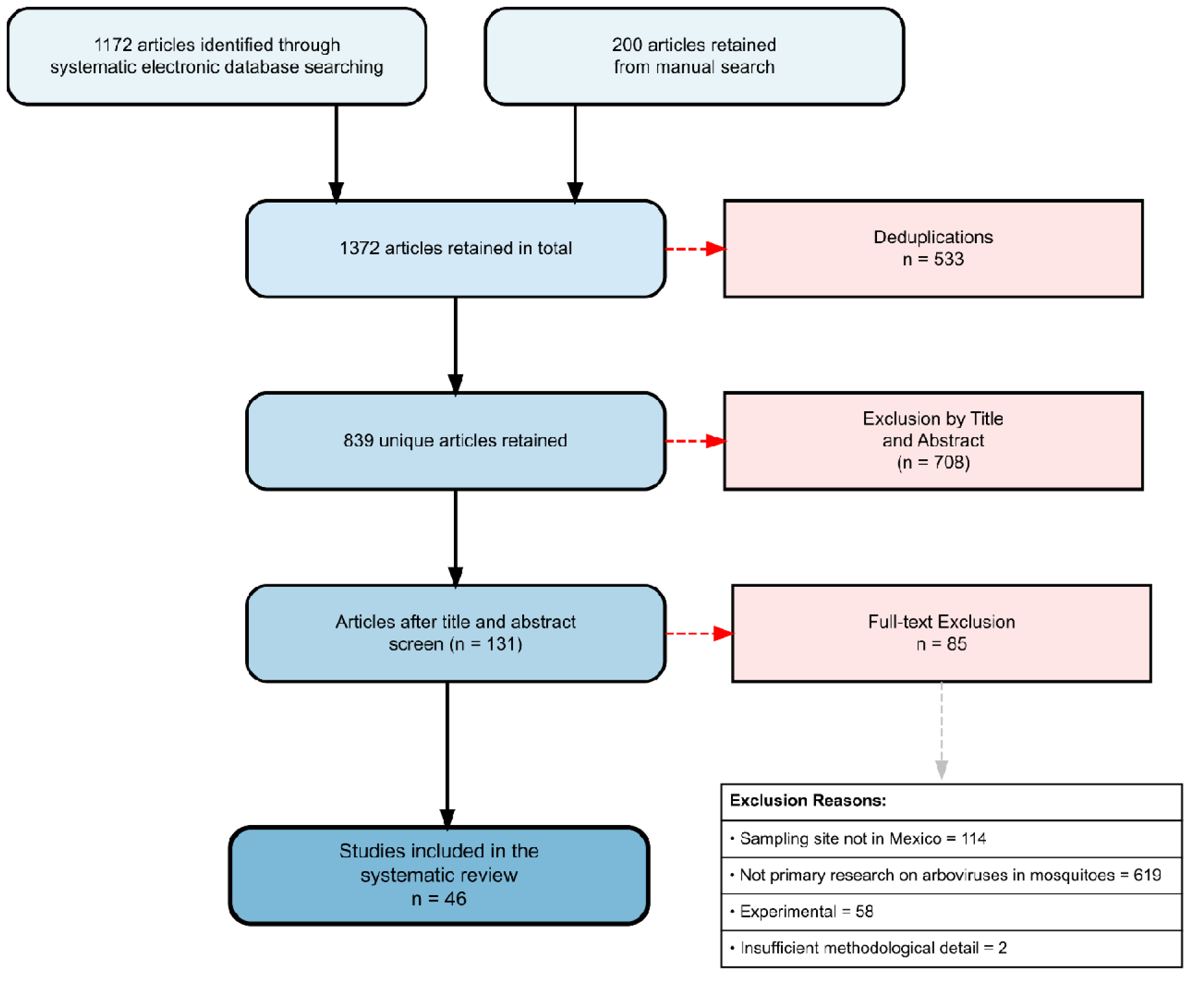
PRISMA diagram of included articles, where “n” represents the number of articles.

### Data collection and synthesis

Post-selection, we extracted spatial and methodological data into Excel for analysis in R (v4.0.1; S1 Table). Synthesis eligibility was then determined by data relevance and completeness. While all selected studies remain in the review, those with insufficient variables were omitted from specific quantitative synthesis subsets; given substantial heterogeneity, as detailed in S1 Table.

### Sampled locations and spatial analysis

Geographic inequality in sampling was quantified using the Gini coefficient (*ineq*) (S3 Fig). Spatial analysis was conducted using curated municipality-level minimum infection rates (MIR). Variability in pool sizes means the MIR serves as a conservative estimate under standard pooled sampling assumptions. Spatial autocorrelation of MIR was evaluated using Global and Local Moran’s I with k-nearest neighbor weights (k=4; *spdep*). Municipalities with at least one positive pool were included. Records lacking sufficient data to compute MIR (i.e., total mosquitoes tested or pool counts) were excluded. In addition, municipalities with extremely low mosquito sample sizes or disproportionate pooling were critically assessed and excluded when MIR values were likely inflated by sampling artefacts (e.g., Chihuahua), thereby ensuring the robustness of spatial inference. Significance was assessed using a Monte Carlo permutation test (999 iterations). Spatial autocorrelation of sampling intensity was evaluated separately. Associations between sampling effort (total pools) and MIR were examined using Pearson and Spearman correlations to assess linear and rank-based relationships (S2 Fig).

### Mosquito-arbovirus associations

#### Mosquito-arbovirus network*s*

Networks and ordination matrix were summarized using pooled detection-based prevalence values across taxa and restricted to vertebrate-pathogenic arboviruses. Bipartite and circular network structures were constructed (*igraph*); node-level degree centrality, species virus richness and virus host breadth were computed. Network edges represent co-detection of arboviruses within mosquito taxa and were treated as binary (presence/absence) due to heterogeneity in reporting across studies. Non-metric multidimensional scaling (NMDS; *vegan*) ordination was based on a species-by-virus prevalence matrix (S7 Table).

#### Species-specific models

Species–specific detection patterns were evaluated using binomial generalized linear mixed models (GLMMs; *lme4*) [14]. The analytical unit was the mosquito pool, coded as positive (1) or negative (0). To reduce instability and control study heterogeneity in model estimates, a minimum sample threshold (<10 mosquitoes) and a study identity fitted as a random intercept were applied:

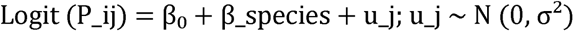

Mosquito species was a fixed effect, with *Aedes aegypti* specified as the reference. Odds ratios (OR) and 95% confidence intervals (CI) were extracted and statistical significance was defined as p ≤ 0.05. Low-frequency taxa were grouped into *Aedes spp* and *Culex spp* categories. We were restricted to ZIKV and vertebrate-pathogenic flavivirus (DENV, ZIKV and WNV) because other virus groups lacked sufficient species-specific pool-level data for a stable estimation (S8 Table).

### Vertical transmission in primary mosquito vectors

Minimum Infection Rate (MIR) was calculated as: (Number of positive pools / Total mosquitoes tested) × 1000. Transmission was classified as vertical (virus detected in immature stages or males) or horizontal (adult females only). Records lacking sufficient data to compute MIR or failing to state mosquito sex and/or life stage were excluded (S9 Table). Chi-square tests (p ≤ 0.05) compared pool positivity between vertical and horizontal transmission studies for DENV, ZIKV, and CHIKV separately [15].

## RESULTS

### Brief study characteristics

The search yielded 839 unique records. All records were screened and 708 were not relevant based on the title and abstract. An additional 85 were excluded based on full-text eligibility. A total of 46 studies were included in this review; however, not all contributed to quantitative synthesis because synthesis-specific eligibility depended on the availability of complete pool, location, taxonomic, or transmission data. These studies comprised Original Articles (36; [3, 4, 7–10, 12, 18, 20, 21, 26–29, 31–34, 39–42, 44, 45, 47–54, 56–59, 61]), Short Communications (4; [36, 37, 46, 60]), Scientific Notes (4; [19, 35, 43, 55]), Dispatches (1; [30]), and Brief Report (1; [38]). A total of 78 mosquito species were screened; 28 species tested positive for arboviruses. The most surveyed species by authors was *Ae. aegypti*, followed by *Culex* (*Cx.*) *quinquefasciatus* and *Cx. nigripalpus* (S1 Table).

### Spatial analyses

Geographic coverage was uneven (pool-level Gini = 0.475) (S3 Fig). MIR spatial autocorrelation was significantly positive (Moran’s I = 0.139; Z = 2.65; p = 0.004; Monte Carlo p = 0.016), showing a non-random spatial distribution. The highest interpretable MIR values were observed in Sierra Monte Negro State Reserve (Morelos; MIR = 68.36), San Marcos (Guerrero; MIR = 57.85), Tlaquepaque (Jalisco; MIR = 55.27), Tapachula (Chiapas; MIR = 28.21) and Ocozocoautla de Espinosa (Chiapas; MIR = 22.22) (Fig. 2). Local Moran’s I did not identify statistically significant municipality-level clusters (Moran’s I = 0.092; p = 0.045; S4 Fig), supporting a diffuse rather than focal spatial structure. The MIR was not positively associated with sampling intensity (Pearson r = −0.188, p = 0.288; S2 Fig) and the rank-based relationship was negative (Spearman ρ = −0.60, p < 0.001; S5 Table), suggesting limited influence of sampling effort on spatial patterns.

**Fig 2.**
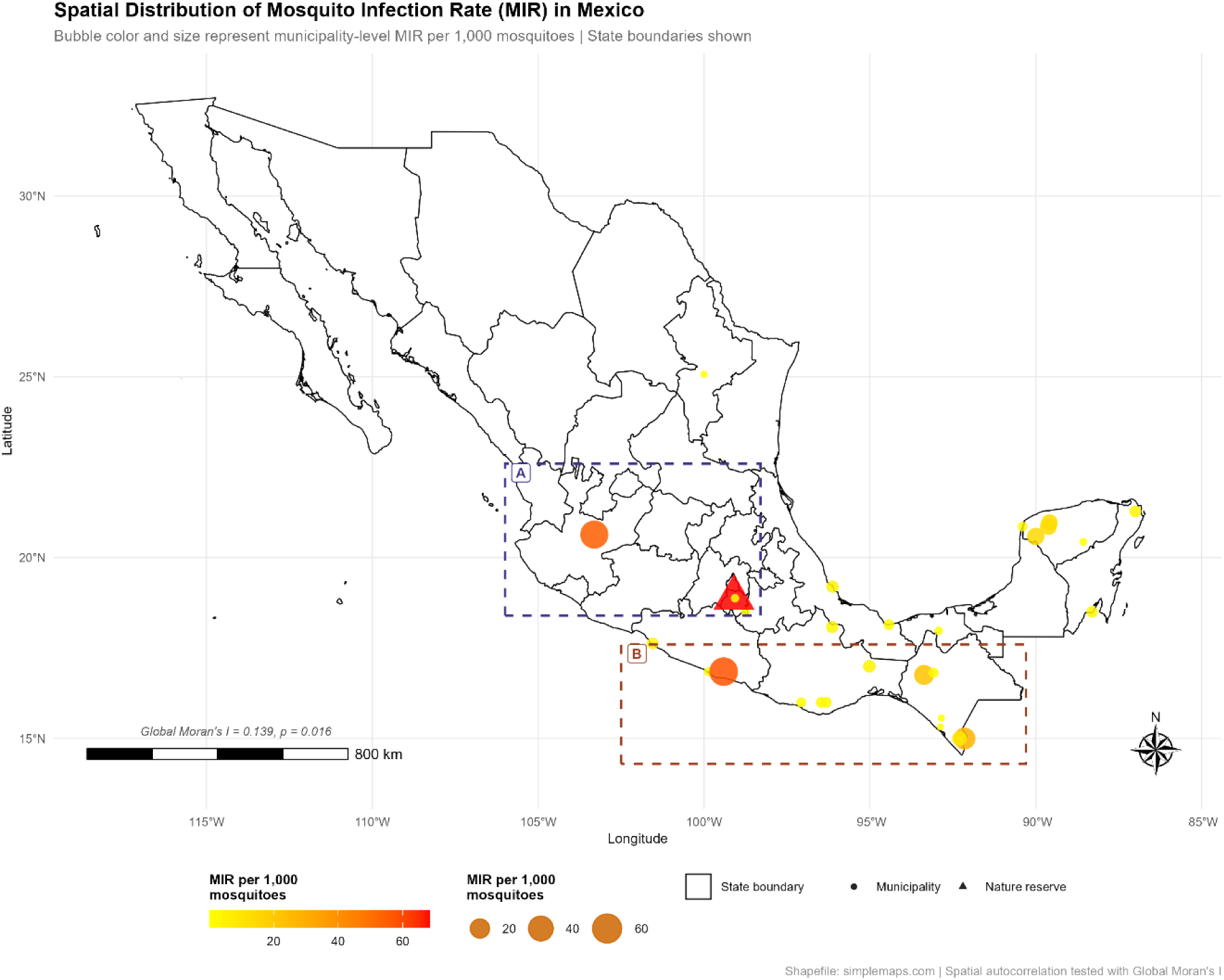
Municipality-level minimum infection rates (MIR) of arboviruses across México. Point size and color gradient represent MIR magnitude. Inset A and B mark the location with elevated MIR in the south-central and southeastern region, respectively. Local Moran’s I did not detect significant clustering, supporting a diffuse spatial structure. Map created in R (version 4.0.1).

### Mosquito–arbovirus associations

#### Mosquito–arbovirus networks

Network analyses revealed non-random associations between mosquitoes and arboviruses by both virus group and mosquito genus. NMDS ordination demonstrated genus-level separation of detection profiles (stress ≈ 0), with *Aedes* and *Culex* genera occupying distinct regions of the reduced ordination space (S6 Fig). The network comprised 11 nodes and 12 edges (connectance = 0.40). *Ae. aegypti* exhibited the highest connectivity (degree = 4); most other species were peripheral (degree ≤ 2; S7 Table). Vertebrate-pathogenic flaviviruses showed generalist behavior across diverse taxa, while orthobunyaviruses remained restricted to *Ae. taeniorhynchus* and *Cx. nigripalpus*. The latter signal, however, lacked cross-study replication, originating from a single study despite high sampling effort (*n* = 371; [53]).

#### Generalized linear mixed models (GLMMs)

GLMM analysis revealed species-specific detection patterns relative to *Ae. aegypti*. Detection of vertebrate-pathogenic flaviviruses was significant in *Cx. quinquefasciatus* (OR = 1.40, p = 0.001) but lower in *Aedes spp.* (OR = 0.18, p = 0.018), with no associations for *Ae. albopictus* (OR = 0.50, p = 0.054) or *Culex spp* (OR = 0.59, p = 0.130). Similarly, ZIKV models identified *Cx. quinquefasciatus* as the only taxon with significantly higher odds of positive pools (OR = 1.88, p < 0.001); *Ae. albopictus*, *Aedes spp.* and *Culex spp*. remained non-significant (Fig. 3; S8 Table). Random intercepts ranged from −3.05 to 2.45, indicating between-study variability, which was accounted for in the model.

**Fig 3.**
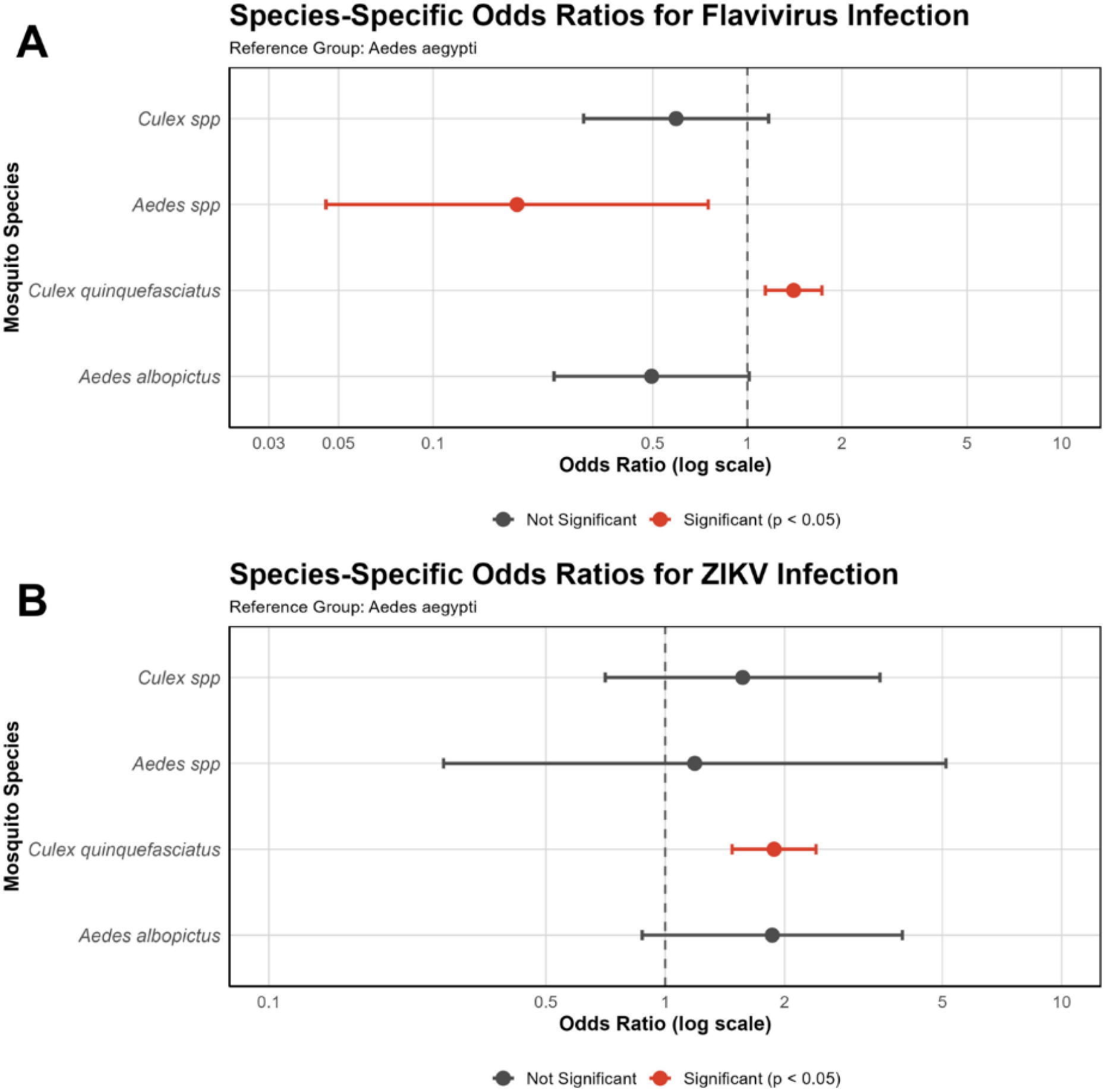
GLMM-derived odds ratios (OR, 95% CI) for arbovirus detection in mosquito taxa relative to *Aedes aegypti*. Panels represent vertebrate-pathogenic flavivirus and ZIKV model fitted using pool-level outcomes; significant associations (p ≤ 0.05) are indicated.

### Vertical transmission in primary mosquito vectors

Studies were classified as horizontal (27; [7–9, 20, 21, 27, 28, 30, 32–35, 37, 39–43, 46–49, 54, 56–58, 61]), vertical (6; [10, 19, 31, 51, 53, 55]), or both (5; [18, 36, 44, 45, 59]) transmission modes (S9 Table). Due to limited data on other species, analysis was restricted to *Ae. aegypti*. Virus-specific analyses revealed heterogeneous patterns. For DENV, vertical transmission studies yielded an MIR of 2.03 (4.75% positivity), compared with an MIR of 4.71 (5.61% positivity) in horizontal studies, with no significant difference (χ² = 1.01, p = 0.315). For ZIKV, vertical transmission showed lower MIR (3.15 vs. 5.15) and significantly fewer positive pools (1.5% vs. 6.0%; χ² = 4.79, p = 0.029), although confidence intervals were wide (OR = 0.247). In contrast, CHIKV exhibited a pronounced disparity, with vertical transmission yielding an MIR of 1.34 (2.03%) positivity versus an MIR of 11.14 (14.52% positivity) in horizontal studies (χ² = 49.47, p < 0.0001, OR = 0.122). The absence of a statistical difference for DENV suggests comparable detection between transmission modes, with no definitive maintenance roles.

## DISCUSSION

Evidence of the circulation of mosquito-borne viruses in México is dispersed, limiting inference on spatial structure and maintenance-relevant associations. This study provides a resolved overview of the structural and spatial dynamics of arbovirus detection patterns across mosquito taxa in México. To achieve this, we reconstruct an analytical framework by consolidating 46 entomo-virological reports spanning nearly two decades.

Spatial analyses demonstrated that arbovirus detections in México are structured but not confined to discrete hotspots. Detection patterns align with historical arbovirus activity in south-central and southeastern México, favoring sustained vector abundance [1, 16, 17]. Elevated values were observed mainly in urban communities: Tlaquepaque (Jalisco), San Marcos (Guerrero), and Tapachula (Chiapas), and fragmented reserve edges and agricultural landscapes: Sierra Monte Negro State Reserve (Morelos) and Ocozocoautla de Espinosa (Chiapas), consistent with ecological descriptions from previous studies [8, 18–21] and aligning with the absence of localized clustering. This landscape profile, though inconsistently classified across studies, reflects ecological transition zones where vegetation and land use interface, a profile increasingly recognized as transmission corridors for the regional expansion of yellow fever in South America [6]. Cautious interpretation is essential, as MIR reflects entomological infection rather than human incidence; however, uneven geographic coverage and sampling inequality underline the need for spatially balanced entomo-virological surveillance.

Species-specific modeling places *Ae. aegypti* at the center of the mosquito–arbovirus network and the primary urban vector in the Americas. However, genus-level separation and cross-genus detections show that viral exposure is structured but not strictly compartmentalized. Vertebrate-pathogenic flaviviruses displayed the widest host breadth, aligning with other regional syntheses [14], whereas orthobunyaviruses were taxonomically restricted. Consequently, our findings represent a simplified structural overview with reduced dimensionality rather than a high-resolution ecological pattern.

This taxonomic breadth is particularly evident in the *Culex* genus. *Cx. quinquefasciatus* retained elevated detection odds for ZIKV and remained significant in vertebrate-pathogenic flavivirus models. This suggests a ZIKV-driven signal rather than a general flavivirus trait. Part of this signal may reflect pooled-detection artefacts, particularly when the abdomens of blood-fed mosquitoes were not analyzed separately, as feeding status was rarely reported across studies. Feeding patterns may inflate apparent ZIKV-associated odds of infection. Importantly, field detection does not equate to competence; experimental studies confirm that *Cx. quinquefasciatus* is refractory to ZIKV infection [22–24]. Instead, its recurrent but peripheral network placement suggests repeated ecological exposure rather than primary transmission. This pattern of maintenance warrants validation and continued investigations under cautious epidemiological interpretation, as supported by predictive models [25].

Vertical transmission analysis reveals virus-specific heterogeneity and methodological inconsistencies. Although vertical infection in *Ae. aegypti* was documented, pooled MIR and positivity were lower compared with horizontal transmission for most virus groups. The absence of a significant difference for DENV, contrasted with disparities for CHIKV and ZIKV, indicates that vertical transmission influence differs amongst key viruses, consistent with Pan-American observations [15]. This deduction supports the hypothesis that vertical transmission may contribute to the maintenance of DENV in México. Limiting the inference is the proportion of studies lacking reports on mosquito sex, developmental stage, or gonotrophic status, emphasizing the need for harmonized reporting standards.

Entomo-virological evidence in México is systematically fragmented, but this synthesis resolves spatial and taxonomic patterns across a fragmented evidence base. As a quantitative synthesis, it reveals an MIR-based spatial structure, virus-specific differences in mosquito pool positivity and a non-exclusive mosquito-arbovirus network. Evidence of vertical transmission is limited and lacks cross-species alignment, suggesting a constrained but virus-specific role in maintenance.

These findings support the need for standardized, multi-taxon monitoring that retains focus on primary vectors while accounting for secondary detection patterns. Strengthening reporting standards, expanding spatial coverage, and experimentally validating field-detected associations will be essential for refined arbovirus surveillance.

## LIMITATIONS

Several limitations should be considered as studies varied in reporting completeness and/or eligibility for this review, but not all quantitative syntheses. Pool-based metrics (e.g., MIR) rather than model-based infection estimators may underestimate true infection prevalence and reduce precision, particularly where pool sizes vary and blood-fed mosquitoes are not distinguished. Species-level sampling was uneven across studies, limiting ecological interpretation across heterogeneous landscapes and network inferences reflect co-detection rather than confirmed transmission pathways. Geographic coverage remains incomplete, with several regions of México underrepresented in published surveillance. These patterns likely reflect a composite of episodic outbreaks rather than a stable spatio-temporal structure. Despite these constraints, the integration of spatial statistics, mixed modeling, and network structure provides a more resolved understanding of arbovirus detection patterns in México than was previously attainable.

## Data Availability

All data produced in the present work are contained in this manuscript and supplementary materials are available upon reasonable request to the authors.

## ACKNOWLEDGMENTS

We appreciate Prof. A.A. Obayomi and Prof. W.F. Sule for their suggestions on statistical models in this manuscript.

## Author contributions

**Conceptualization:** Ayokunle D. Obayomi

**Data curation:** Ayokunle D. Obayomi, Marvellous Asubiojo, Jonathan P. Cisneros.

**Formal analysis:** Ayokunle D. Obayomi, Marvellous Asubiojo, Omar Domínguez-Acosta.

**Methodology:** Ayokunle D. Obayomi, Marvellous Asubiojo, Omar Domínguez-Acosta.

**Supervision:** Ma Isabel Salazar.

**Writing-original draft:** Ayokunle D. Obayomi, Jonathan P. Cisneros.

**Writing – review & editing:** Ayokunle D. Obayomi, Temidayo O. Elufisan, Ma Isabel Salazar.

## Data Availability Statement

All relevant data underlying the findings of this study are included within the manuscript and supporting information files.

## Supporting information

**S1 Table. Master extraction dataset of entomo-virological studies included in the review**

**S2 Fig. Correlations between sampling effort and minimum infection rate**

**S3 Fig. Lorenz curve showing inequality in the geographic distribution**

**S4 Fig. Local Moran’s I scatterplot**

**S5 Table. Municipality-level spatial analysis dataset**

**S6 Fig. Mosquito–arbovirus network structure and NMDS ordination**

**S7 Table. Mosquito–arbovirus association matrix and network metrics**

**S8 Table. Datasets and model outputs for GLMM analyses**

S9 Table. Virus-specific datasets for vertical transmission analyses in *Aedes aegypti*

**S10 Table. PRISMA 2020 checklist**

## REFERENCES

1. Gutierrez B, da Silva Candido D, Bajaj S, Rodriguez Maldonado AP, Ayala FG, Torre Rodriguez ML, et al. Convergent trends and spatiotemporal patterns of Aedes-borne arboviruses in México and Central America. PLoS Negl Trop Dis. 2023;17(9):e0011169. 10.1371/journal.pntd.0011169 PMID: 37729647 PMCID: PMC10515158.

2. World Health Organization (2020). WHO Fact Sheet: Vector-borne diseases. Available online at: http://www.who.int/mediacentre/factsheets/fs387/en/ (Accessed June 10, 2024).

3. Hernández-Villegas EN, Castelán-Sánchez HG, Moreira-Soto A, Vigueras-Galván AL, Jiménez-Rico MA, Rico-Chávez O, et al. Characterization of the Virome in Mosquitoes Across Distinct Habitats in the Yucatán Peninsula, Mexico. Viruses. 2025;17(6):758. doi: 10.3390/v17060758. PMID: 39515717. PMCID: PMC11515717.

4. Hernández-Triana LM, Garza-Hernández JA, Ortega Morales AI, Prosser SWJ, Hebert PDN, Nikolova NI, et al. An Integrated Molecular Approach to Untangling Host–Vector–Pathogen Interactions in Mosquitoes (Diptera: Culicidae) From Sylvan Communities in Mexico. Front Vet Sci. 2021;7:564791. doi: 10.3389/fvets.2020.564791. PMID: 33778018. PMCID: PMC7987826.

5. Rodríguez-González S, Izquierdo-Suzán M, Rocha-Ortega M, Córdoba-Aguilar A. Vector mosquito distribution and richness are predicted by socio-economic and ecological variables. Acta Trop. 2024; 254:107–179. 10.1016/j.actatropica.2024.107179

6. Pan American Health Organization (2026). PAHO: Epidemiological Alert Yellow fever in the Americas Region. Available online: https://www.paho.org/sites/default/files/2026/03/2026-march-13-phe-epidemiological-alert-yellow-fever-final.

7. Correa -Morales F, González-Acosta C, Mejía-Zúñiga D, Huerta H, Pérez-Rentería C, Vázquez-Pichardo M, et al. Surveillance for Zika in Mexico: naturally infected mosquitoes in urban and semi-urban areas. Pathog Glob Health. 2019;113(7):309–14. doi: 10.1080/20477724.2019.1706291. PMID: 31868142. PMCID: PMC6996655.

8. Izquierdo-Suzán M, Zavala-Guerrero PB, Mendoza H, Portela Salomão R, Vázquez-Pichardo M, Von Thaden JJ, et al. Mosquito (Diptera: Culicidae) diversity and arbovirus detection across an urban and agricultural landscape. Acta Trop. 2024;257:107321. doi: 10.1016/j.actatropica.2024.107321. PMID: 38971448.

9. Kirstein OD, Ayora-Talavera G, Koyoc-Cardeña E, Chan Espinoza D, Che-Mendoza A, Cohuo-Rodriguez A, et al. Natural arbovirus infection rate and detectability of indoor female Aedes aegypti from Mérida, Yucatán, Mexico. PLoS Negl Trop Dis. 2021;15(1):e0008972. doi: 10.1371/journal.pntd.0008972. PMID: 33395431. PMCID: PMC7808625.

10. Kirstein OD, Ayora Talavera G, Wei Z, Ciau-Carrilo KJ, Koyoc-Cardeña E, Puerta-Guardo H, et al. Natural Aedes-Borne Virus Infection Detected in Male Adult Aedes aegypti (Diptera: Culicidae) Collected From Urban Settings in Mérida, Yucatán, México. J Med Entomol. 2022;59(4):1336–46. doi: 10.1093/jme/tjac048. PMID: 35536551. PMCID: PMC9279543.

11. Talaga S, Le Goff G, Arana-Guardia R, Baak-Baak CM, García-Rejón JE, García-Suárez O, et al. The mosquitoes (Diptera: Culicidae) of the Mexican Yucatan Peninsula: a comprehensive review on the use of taxonomic names. J Med Entomol. 2024;61(2):274–308. 10.1093/jme/tjad168

12. Dzul-Manzanilla F, Correa-Morales F, Che-Mendoza A, Palacio-Vargas J, Sánchez-Tejeda G, González-Roldan JF, et al. Identifying urban hotspots of dengue, chikungunya, and Zika transmission in Mexico to support risk stratification efforts: a spatial analysis. Lancet Planet Health. 2021;5(5):e277–e285. 10.1016/S2542-5196(21)00030- PMID: 33991427.

13. Shamseer L, Moher D, Clarke M, Ghersi D, Liberati A, Petticrew M, et al. Preferred reporting items for systematic review and meta-analysis protocols (PRISMA-P) 2015: elaboration and explanation. BMJ. 2015; 350:g7647. 10.1136/bmj.g7647 PMID: 25555855

14. McMillan JR, Armstrong PM, Andreadis TG. Patterns of mosquito and arbovirus community composition and ecological indexes of arboviral risk in the northeast United States. PLoS Negl Trop Dis. 2020;14(2): e0008066. 10.1371/journal.pntd.0008066

15. Garcia-Rejon JE, Navarro JC, Cigarroa-Toledo N, Baak-Baak CM. An updated review of the invasive Aedes albopictus in the Americas; geographical distribution, host feeding patterns, arbovirus infection, and the potential for vertical transmission of dengue virus. Insects. 2021;12(11):967. 10.3390/insects12110967 PMID: 34821958 PMCID: PMC8624853

16. Bond JG, Moo-Llanes DA, Ortega-Morales AI, Marina CF, Casas-Martínez M, Danis-Lozano R. Diversity and potential distribution of culicids of medical importance of the Yucatan Peninsula, México. Salud Pública Mex. 2020;62(4):379–387. 10.21149/11208 PMID: 32701983

17. Baak-Baak CM, Moo-Llanes DA, Cigarroa-Toledo N, Puerto FI, Machain-Williams C, Reyes-Solis G, et al. Ecological niche model for predicting distribution of disease-vector mosquitoes in Yucatán State, México. J Med Entomol. 2017;54(4):854–861. 10.1093/jme/tjw243 PMID: 28399230

18. Elizondo-Quiroga D, Medina-Sánchez A, Sánchez-González JM, Eckert KA, Villalobos-Sánchez E, Navarro-Zúñiga AR, et al. Zika Virus in salivary glands of five different species of wild-caught mosquitoes from Mexico. Sci Rep. 2018;8(1):809. doi: 10.1038/s41598-017-18682-3. PMID: 29339773. PMCID: PMC5770356.

19. Ponce-Garcia G, Arredondo-Jiménez JI, Machain-Williams C, Baak-Baak CM, Cigarroa-Toledo N, Reyes-Solis GC, et al. Natural Chikungunya Virus Infection in Larvae and Adult Aedes aegypti From San Marcos, Guerrero, Mexico. J Am Mosq Control Assoc. 2018;34(2):147–50. doi: 10.2987/18-6725.1. PMID: 30044709.

20. Díaz-González EE, Kautz TF, Dorantes-Delgado A, Malo-García IR, Laguna-Aguilar M, Langsjoen RM, et al. First Report of Aedes aegypti Transmission of Chikungunya Virus in the Americas. Am J Trop Med Hyg. 2015;93(6):1325–9. doi: 10.4269/ajtmh.15-0450. PMID: 26416110. PMCID: PMC4674251.

21. Zhou LH, Valdez F, Lopez Gonzalez I, Freysser Urbina W, Ocaña A, Tapia C, et al. Vesicular Stomatitis Virus Transmission Dynamics Within Its Endemic Range in Chiapas, Mexico. Viruses. 2024;16(11):1742. doi: 10.3390/v16111742. PMID: 39598282. PMCID: PMC11598282.

22. Fernandes RS, Campos SS, Ribeiro PS, Raphael L, Bonaldo MC, Lourenço-de-Oliveira R. Culex quinquefasciatus from areas with the highest incidence of microcephaly associated with Zika virus infections in the Northeast Region of Brazil are refractory to the virus. Mem Inst Oswaldo Cruz. 2017;112:577–579. 10.1590/0074-02760170145 PMID: 28793059.

23. Hart CE, Roundy CM, Azar SR, Huang JH, Yun R, Reynolds E, et al. Zika virus vector competency of mosquitoes, Gulf Coast, United States. Emerg Infect Dis. 2017;23(3):559. 10.3201/eid2303.161636 PMID: 28221132 PMCID: PMC5382786.

24. Kenney JL, Romo H, Duggal NK, Tzeng WP, Burkhalter KL, Brault AC, et al. Transmission incompetence of Culex quinquefasciatus and Culex pipiens pipiens from North America for Zika virus. Am J Trop Med Hyg. 2017;96(5):1235. 10.4269/ajtmh.16-0865 PMID: 28419118 PMCID: PMC5417217.

25. Evans MV, Dallas TA, Han BA, Murdock CC, Drake JM. Data-driven identification of potential Zika virus vectors. eLife. 2017;6:e22053. 10.7554/eLife.22053 PMID: 28362220 PMCID: PMC5382534.

26. Apodaca-Medina AI, Torres-Avendaño JI, Rendón-Maldonado JG, Torres-Montoya EH, Flores-López BA, Del Angel RM, et al. First Evidence of Vertical Infection of Dengue Virus 2 in Aedes aegypti Mosquitoes from Sinaloa, Mexico. Vector Borne Zoonotic Dis. 2018;18(4):231–3. doi: 10.1089/vbz.2017.2202. PMID: 29558319.

27. Argaez-Sierra DG, Baak-Baak CM, Garcia-Rejon JE, Cetina-Trejo RC, Tzuc-Dzul JC, Acosta-Viana KY, et al. Entomo-virological surveillance of Flavivirus in mosquitoes in Yucatan State, Mexico. Rev Inst Med Trop Sao Paulo. 2024;66:e56. doi: 10.1590/S1678-9946202466056. PMID: 39111005. PMCID: PMC11302882.

28. Baak-Baak CM, Cigarroa-Toledo N, Pech-May A, Cruz-Escalona GA, Cetina-Trejo RC, Tzuc-Dzul JC, et al. Entomological and virological surveillance for dengue virus in churches in Merida, Mexico. Rev Inst Med Trop Sao Paulo. 2019;61:e9. doi: 10.1590/S1678-9946201961009. PMID: 30726330. PMCID: PMC6362675.

29. Charles J, Tangudu CS, Hurt SL, Tumescheit C, Firth AE, Garcia-Rejon JE, et al. Detection of novel and recognized RNA viruses in mosquitoes from the Yucatan Peninsula of Mexico using metagenomics and characterization of their in vitro host ranges. J Gen Virol. 2018;99(12):1729–38. doi: 10.1099/jgv.0.001157. PMID: 30300283. PMCID: PMC6339736.

30. Cigarroa-Toledo N, Blitvich BJ, Cetina-Trejo RC, Talavera-Aguilar LG, Baak-Baak CM, Torres-Chablé OM, et al. Chikungunya Virus in Febrile Humans and Aedes aegypti Mosquitoes, Yucatan, Mexico. Emerg Infect Dis. 2016;22(10):1804–7. doi: 10.3201/eid2210.152087. PMID: 27648777. PMCID: PMC5038421.

31. Danis-Lozano R, Díaz-González EE, Malo-García IR, Rodríguez MH, Ramos-Castañeda J, Juárez-Zúñiga S, et al. Vertical transmission of dengue virus in Aedes aegypti and its role in the epidemiology of dengue in Mexico. Trop Med Int Health. 2019;24(11):1311–9. doi: 10.1111/tmi.13306. PMID: 31448555.

32. De la Mora-Covarrubias A, Jiménez-Vega F, Treviño-Aguilar SM. Distribución geoespacial y detección del virus del dengue en mosquitos Aedes (Stegomyia) aegypti de Ciudad Juárez, Chihuahua, México. Salud Publica Mex. 2010;52(2):127–33. PMID: 20485869.

33. Deardorff ER, Estrada-Franco JG, Freier JE, Navarro-Lopez R, Da Rosa AT, Tesh RB, et al. Candidate Vectors and Rodent Hosts of Venezuelan Equine Encephalitis Virus, Chiapas, 2006–2007. Am J Trop Med Hyg. 2011;85(6):1146–53. doi: 10.4269/ajtmh.2011.11-0094. PMID: 22144458. PMCID: PMC3225166.

34. Díaz-Quiñonez JA, López-Martínez I, Torres-Longoria B, Vázquez-Pichardo M, Cruz-Ramírez E, Ramírez-González JE, et al. Evidence of the presence of the Zika virus in Mexico since early 2015. Virus Genes. 2016;52(6):855–7. doi: 10.1007/s11262-016-1384-0. PMID: 27557815.

35. Dzul-Manzanilla F, Martínez NE, Cruz-Nolasco M, Gutiérrez-Castro C, López-Damián L, Ibarra-López J, et al. Arbovirus Surveillance and First Report of Chikungunya Virus in Wild Populations of Aedes aegypti from Guerrero, Mexico. J Am Mosq Control Assoc. 2015;31(3):275–7. doi: 10.2987/moco-31-03-275-277.1. PMID: 26375910.

36. Dzul-Manzanilla F, Martínez NE, Cruz-Nolasco M, Gutiérrez-Castro C, López-Damián L, Ibarra-López J, et al. Evidence of vertical transmission and co-circulation of chikungunya and dengue viruses in field populations of Aedes aegypti (L.) from Guerrero, Mexico. Trans R Soc Trop Med Hyg. 2016;110(2):141–4. doi: 10.1093/trstmh/trv106. PMID: 26711697.

37. Eisen L, García-Rejón JE, Gómez-Carro S, Nájera Vázquez MR, Keefe TJ, Beaty BJ, et al. Temporal Correlations Between Mosquito-Based Dengue Virus Surveillance Measures or Indoor Mosquito Abundance and Dengue Case Numbers in Mérida City, México. J Med Entomol. 2014;51(4):885–90. doi: 10.1603/ME14008. PMID: 25118423. PMCID: PMC4273873.

38. Espinoza-Gómez F, López-Lemus AU, Rodriguez-Sanchez IP, Martinez-Fierro ML, Newton-Sánchez OA, Chávez-Flores E, et al. Detection of sequences from a potentially novel strain of cell fusing agent virus in Mexican Stegomyia (Aedes) aegypti mosquitoes. Arch Virol. 2011;156(7):1263–7. doi: 10.1007/s00705-011-0967-2. PMID: 21409540.

39. Estrada-Franco JG, Fernández-Santos NA, Adebiyi AA, López-López MJ, Aguilar-Durán JA, Hernández-Triana LM, et al. Vertebrate-Aedes aegypti and Culex quinquefasciatus (Diptera)-arbovirus transmission networks: Non-human feeding revealed by meta-barcoding and next-generation sequencing. PLoS Negl Trop Dis. 2020;14(12):e0008867. doi: 10.1371/journal.pntd.0008867. PMID: 33382710. PMCID: PMC7808726.

40. Farfan-Ale JA, Loroño-Pino MA, Garcia-Rejon JE, Hovav E, Powers AM, Lin M, et al. Detection of RNA from a Novel West Nile-like Virus and High Prevalence of an Insect-specific Flavivirus in Mosquitoes in the Yucatan Peninsula of Mexico. Am J Trop Med Hyg. 2009;80(1):85–95. doi: 10.4269/ajtmh.2009.80.85. PMID: 19141845. PMCID: PMC2733075.

41. Farfan-Ale JA, Loroño-Pino MA, Garcia-Rejon JE, Soto V, Lin M, Staley M, et al. Detection of Flaviviruses and Orthobunyaviruses in Mosquitoes in the Yucatan Peninsula of Mexico in 2008. Vector Borne Zoonotic Dis. 2010;10(8):777–83. doi: 10.1089/vbz.2009.0196. PMID: 20497010.

42. García-Rejón JE, Loroño-Pino MA, Farfán-Ale JA, Flores-Flores LF, López-Uribe MP, Najera-Vazquez MR, et al. Mosquito Infestation and Dengue Virus Infection in Aedes aegypti Females in Schools in Mérida, México. Am J Trop Med Hyg. 2011;84(3):489–96. doi: 10.4269/ajtmh.2011.10-0654. PMID: 21363991. PMCID: PMC3042828.

43. García-Rejón JE, Ulloa-Garcia A, Cigarroa-Toledo N, Machain-Williams C, Cetina-Trejo RC, Talavera-Aguilar LG, et al. Determining the mosquito (Diptera: Culicidae) fauna and identifying alphaviruses in mangroves of the Yucatan Coast, Mexico. J Am Mosq Control Assoc. 2023;39(2):134–7. doi: 10.2987/22-7080. PMID: 37376043.

44. García-Rejón JE, Ulloa-Garcia A, Cigarroa-Toledo N, Pech-May A, Machain-Williams C, Cetina-Trejo RC, et al. Study of Aedes aegypti population with emphasis on the gonotrophic cycle length and identification of arboviruses: implications for vector management in cemeteries. Rev Inst Med Trop Sao Paulo. 2018;60:e44. doi: 10.1590/S1678-9946201860044. PMID: 30156637. PMCID: PMC6111166.

45. Guerbois M, Fernandez-Salas I, Azar SR, Danis-Lozano R, Alpuche-Aranda CM, Leal G, et al. Outbreak of Zika Virus Infection, Chiapas State, Mexico, 2015, and First Confirmed Transmission by Aedes aegypti Mosquitoes in the Americas. J Infect Dis. 2016;214(9):1349–56. doi: 10.1093/infdis/jiw210. PMID: 27227829. PMCID: PMC5079361.

46. Günther J, Martínez-Muñoz JP, Pérez-Ishiwara DG, Salas-Benito J. Evidence of Vertical Transmission of Dengue Virus in Two Endemic Localities in the State of Oaxaca, Mexico. Intervirology. 2007;50(5):347–52. doi: 10.1159/000107272. PMID: 17684345.

47. Hernández-Acosta E, De la Mora Covarrubias A, Fernández-Salas I, Escárcega-Ávila A, Passalacqua Olivera I, Jiménez-Vega F. Detection of chikungunya virus in Aedes aegypti in Ciudad Juárez, Chihuahua, Mexico. J Vector Borne Dis. 2025;62:228–32.

48. Hidalgo-Martínez A, Puerto FI, Farfán-Ale JA, García-Rejón JE, Rosado-Paredes EP, Méndez-Galván J, et al. Prevalencia de infección por el virus del Nilo occidental en dos zoológicos del estado de Tabasco. Salud Publica Mex. 2008;50(1):76–85. PMID: 18373024.

49. Huerta H, González-Roldán JF, Sánchez-Tejeda G, Correa-Morales F, Romero-Contreras FE, Cárdenas-Flores R, et al. Detection of Zika virus in Aedes mosquitoes from Mexico. Trans R Soc Trop Med Hyg. 2017;111(7):328–31. doi: 10.1093/trstmh/trx056. PMID: 29016911. PMCID: PMC5759163.

50. Ibarra-Juarez L, Eisen L, Bolling BG, Beaty BJ, Blitvich BJ, Sanchez-Casas RM, et al. Detection of West Nile virus-specific antibodies and nucleic acid in horses and mosquitoes, respectively, in Nuevo Leon State, northern Mexico, 2006–2007. Med Vet Entomol. 2012;26(4):441–6. doi: 10.1111/j.1365-2915.2012.01014.x. PMID: 22449103.

51. Izquierdo-Suzán M, Zárate S, Torres-Flores J, Correa-Morales F, González-Acosta C, Sevilla-Reyes EE, et al. Natural Vertical Transmission of Zika Virus in Larval Aedes aegypti Populations, Morelos, Mexico. Emerg Infect Dis. 2019;25(8):1477–84. doi: 10.3201/eid2508.181533. PMID: 31310214. PMCID: PMC6649326.

52. Kopp A, Gillespie TR, Hobelsberger D, Estrada A, Harper JM, Miller RA, et al. Provenance and Geographic Spread of St. Louis Encephalitis Virus. mBio. 2013;4(3):e00322–13. doi: 10.1128/mBio.00322-13. PMID: 23760460. PMCID: PMC3684824.

53. Kopp A, Hübner A, Zirkel F, Hobelsberger D, Estrada A, Jordan I, et al. Detection of Two Highly Diverse Peribunyaviruses in Mosquitoes from Palenque, Mexico. Viruses. 2019;11(9):832. doi: 10.3390/v11090832. PMID: 31500331. PMCID: PMC6784013.

54. Lopez-Apodaca LI, Zarza H, Zamudio-Moreno E, Nuñez-Avellaneda D, Baak-Baak CM, Reyes-Solis GC, et al. Molecular survey of Zika virus in the animal-human interface in traditional farming. Front Vet Sci. 2022;9:1057686. doi: 10.3389/fvets.2022.1057686. PMID: 36505717. PMCID: PMC9731998.

55. Martínez NE, Dzul-Manzanilla F, Gutiérrez-Castro C, Ibarra-López J, Bibiano-Marín W, López-Damián L, et al. Natural Vertical Transmission of Dengue-1 Virus in Aedes aegypti Populations in Acapulco, Mexico. J Am Mosq Control Assoc. 2014;30(2):143–6. doi: 10.2987/14-6402.1. PMID: 25055562.

56. Méndez-Galván J, Sánchez-Casas RM, Gaitan-Burns A, Díaz-González EE, Ibarra-Juarez LA, Medina de la Garza CE, et al. Detection of Aedes aegypti Mosquitoes Infected with Dengue Virus as a Complementary Method for Increasing the Sensitivity of Surveillance: Identification of Serotypes 1, 2, and 4 by RT-PCR in Quintana Roo, Mexico. Southwest Entomol. 2014;39(2):307–16. doi: 10.3958/059.039.0208.

57. Nuñez-Avellaneda D, Cetina-Trejo RC, Zamudio-Moreno E, Baak-Baak C, Cigarroa-Toledo N, Reyes-Solis G, et al. Evidence of Zika Virus Infection in Pigs and Mosquitoes, Mexico. Emerg Infect Dis. 2021;27(2):574–7. doi: 10.3201/eid2702.201452. PMID: 33496554. PMCID: PMC7853569.

58. Sanchez-Casas RM, Alpuche-Delgado RH, Blitvich BJ, Diaz-Gonzalez EE, Ramirez-Jimenez R, Zarate-Nahon EA, et al. Detection of Dengue Virus Serotype 2 in Aedes aegypti in Quintana Roo, Mexico, 2011. Southwest Entomol. 2013;38(1):109–17. doi: 10.3958/059.038.0115.

59. Sanchez-Rodríguez OS, Sanchez-Casas RM, Laguna-Aguilar M, Alvarado-Moreno MS, Zarate-Nahon EA, Ramirez-Jimenez R, et al. Natural Transmission of Dengue Virus by Aedes albopictus at Monterrey, Northeastern Mexico. Southwest Entomol. 2014;39(3):459–68. doi: 10.3958/059.039.0307.

60. Sotomayor-Bonilla J, Abella-Medrano CA, Chaves A, Álvarez-Mendizábal P, Rico-Chávez Ó, Ibáñez-Bernal S, et al. Potential Sympatric Vectors and Mammalian Hosts of Venezuelan Equine Encephalitis Virus in Southern Mexico. J Wildl Dis. 2017;53(3):657–61. doi: 10.7589/2016-11-249. PMID: 28350212.

61. Ulloa A, Ferguson HH, Méndez-Sánchez JD, Danis-Lozano R, Casas-Martínez M, Bond JG, et al. West Nile Virus Activity in Mosquitoes and Domestic Animals in Chiapas, México. Vector Borne Zoonotic Dis. 2009;9(5):555–60. doi: 10.1089/vbz.2008.0087. PMID: 19442013.

