## Supplementary figures and images for "Structural and spatial dynamics of mosquito-arbovirus associations in México (2007 to 2025): A systematic review and quantitative synthesis"

### S2 Fig.pdf

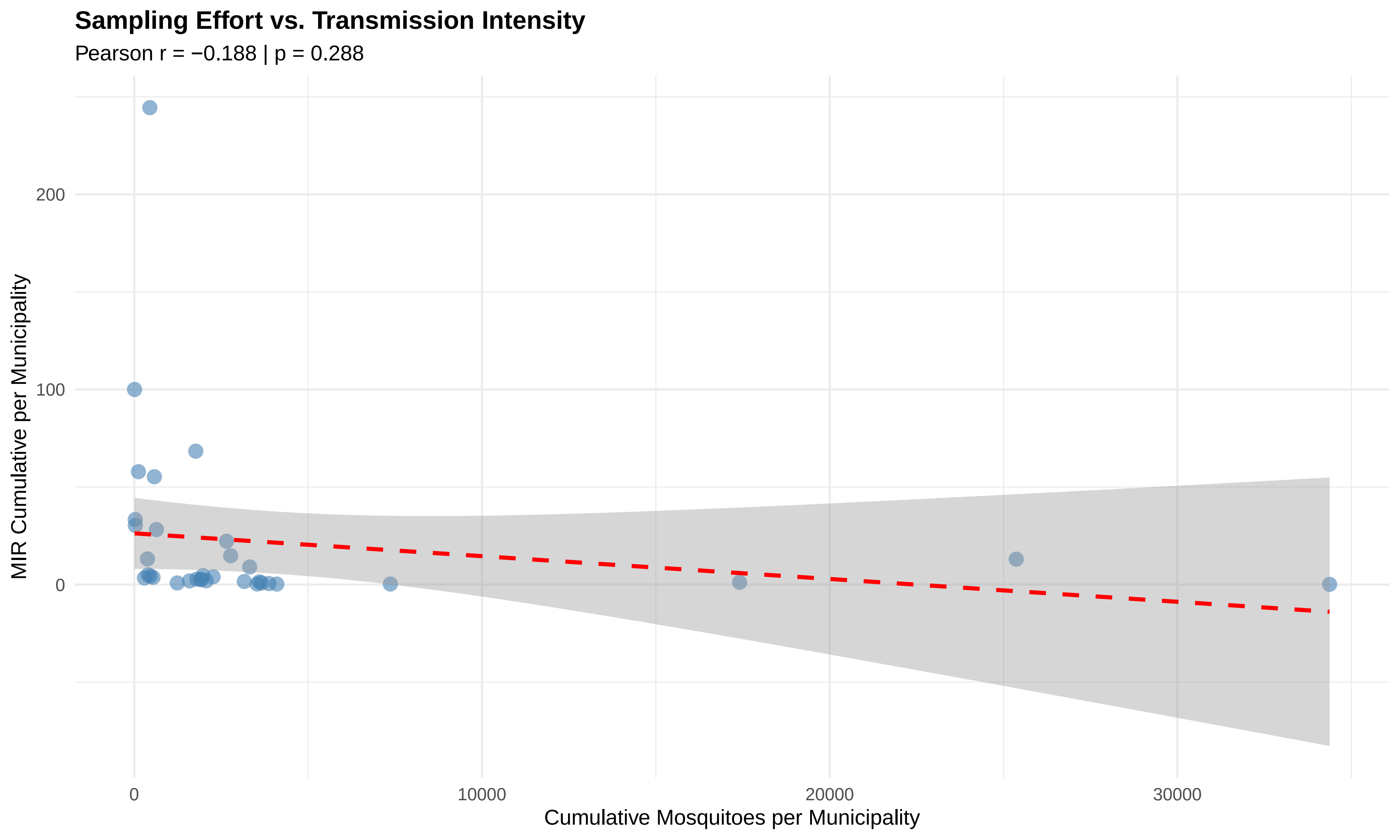

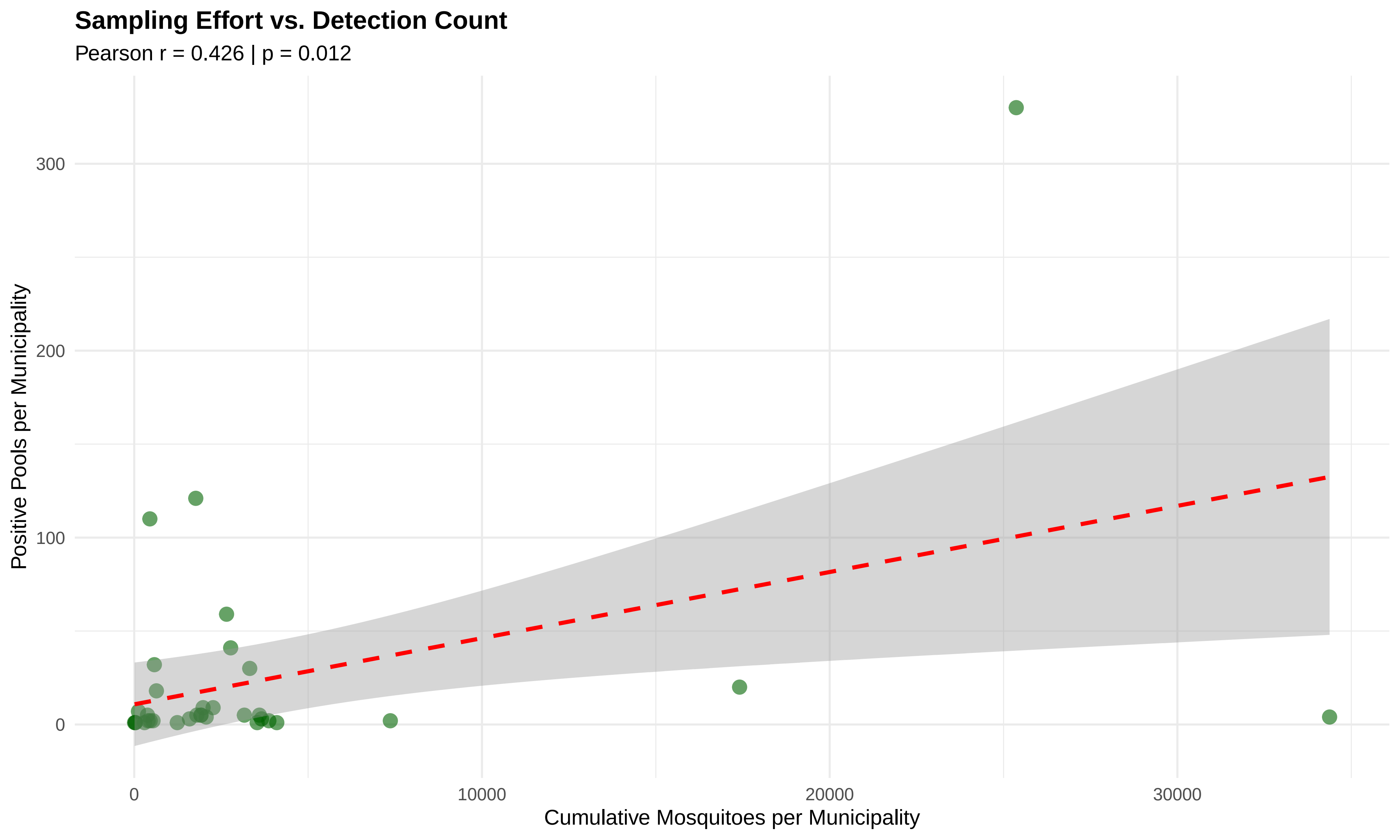

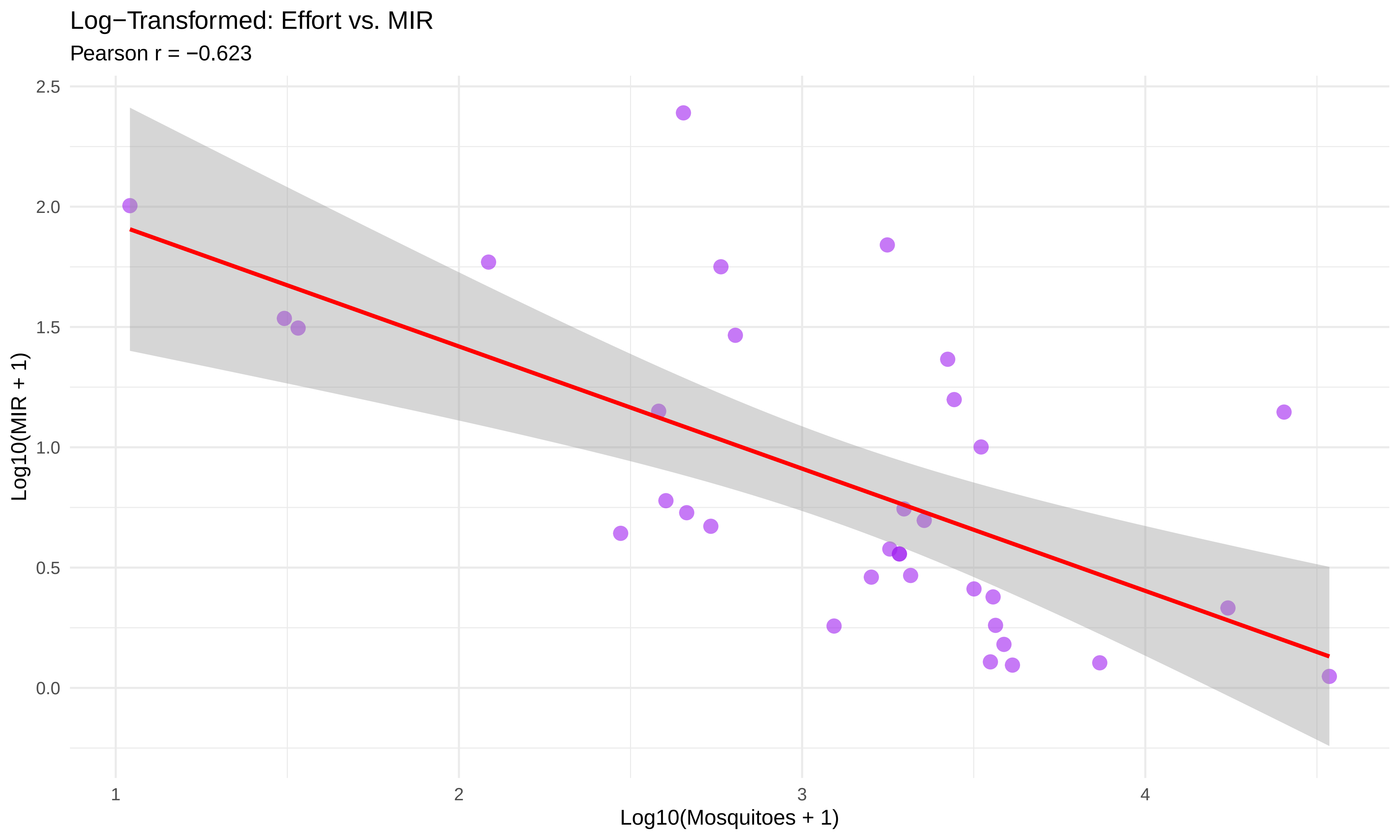

### S3 Fig.pdf

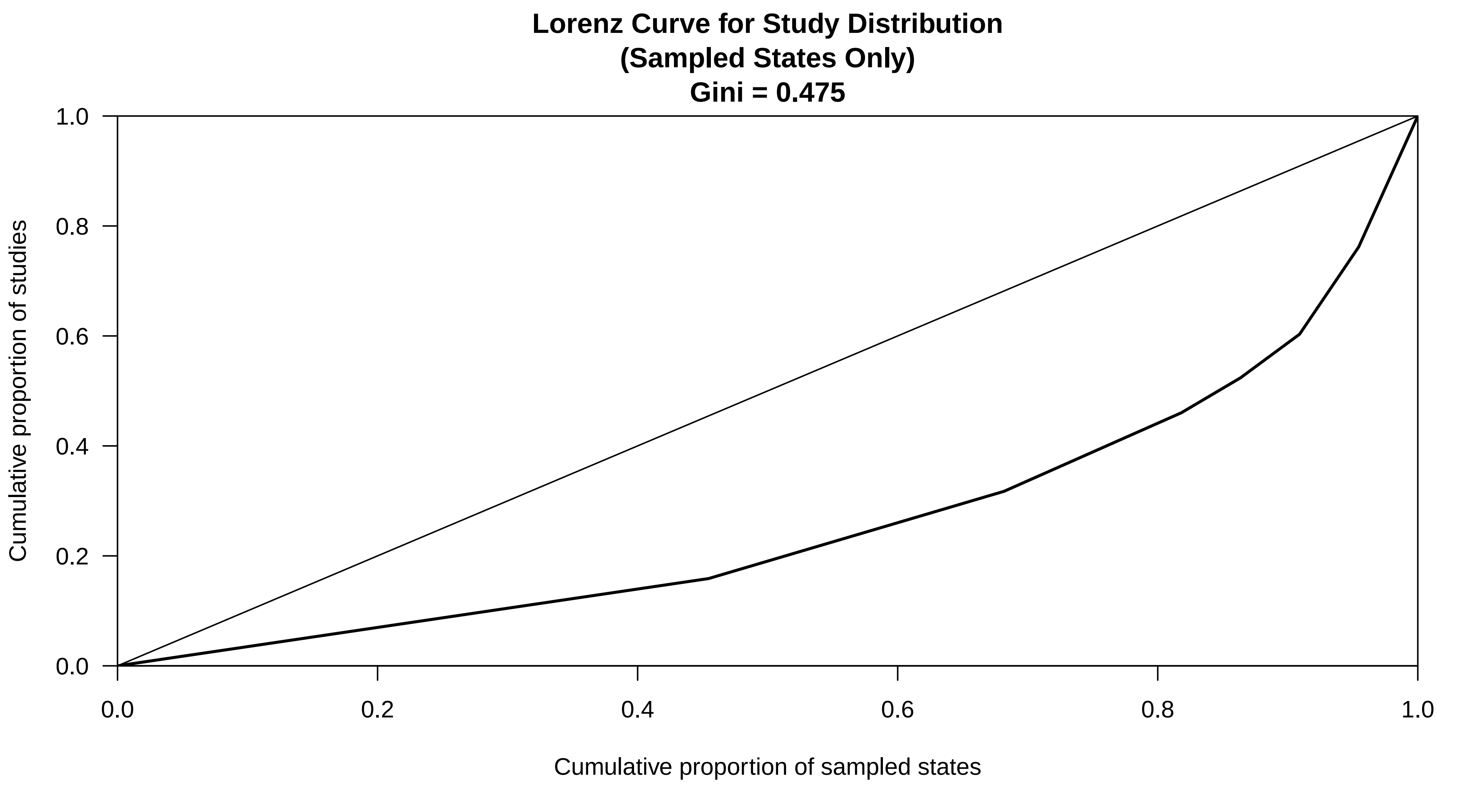

### S4 Fig.pdf

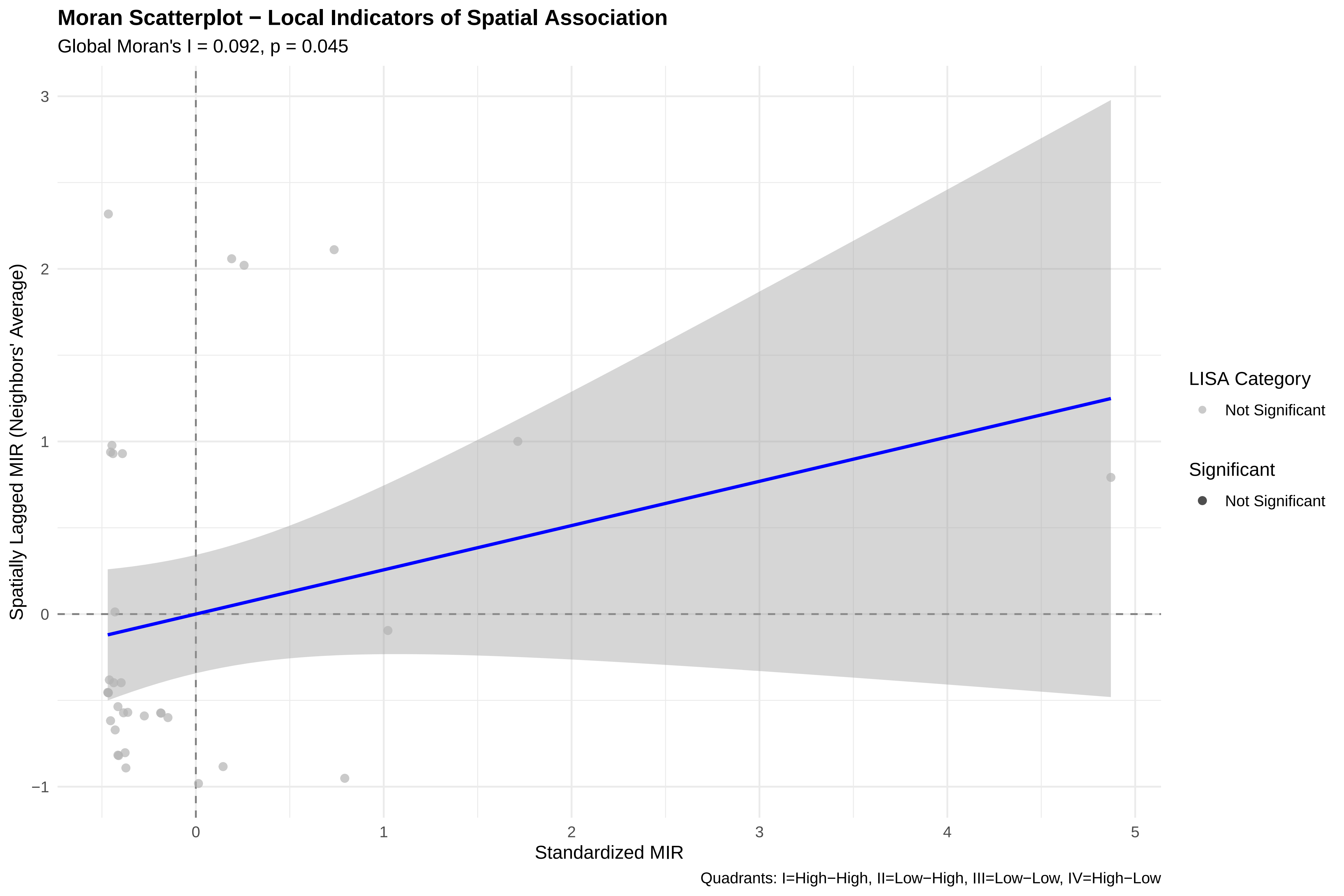

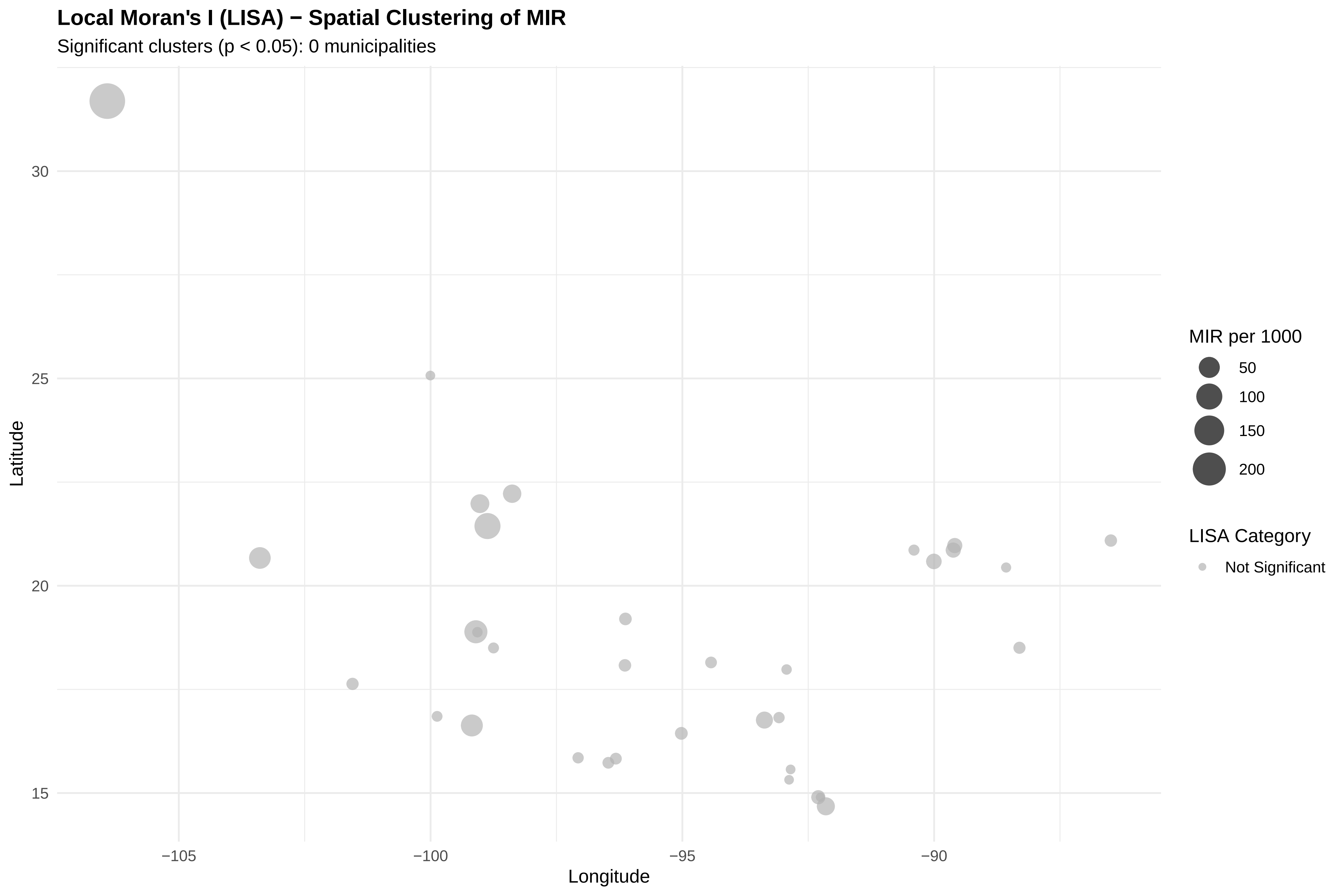

### S6 Fig.png

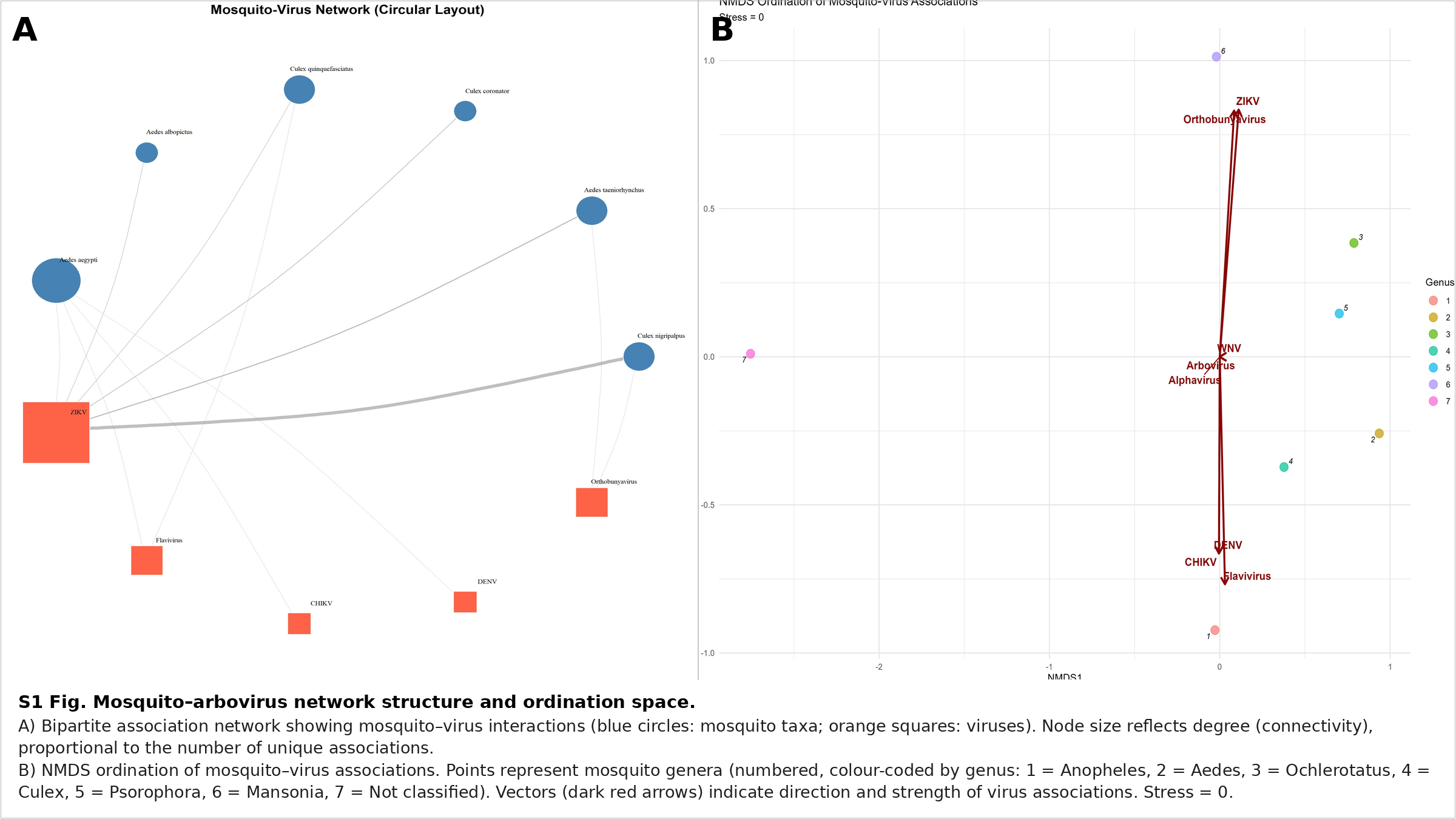
